# Barriers and Enablers to Cervical Cancer Screening Among Culturally Diverse Women in Australia: A Mixed Methods Study

**DOI:** 10.1101/2025.09.02.25334492

**Authors:** Daniella Edward

**Affiliations:** School of Public Health and Medicine, Adelaide University

## Abstract

**Background and Aims:** Despite advances in cervical cancer prevention through HPV vaccination and self-collection screening options, participation remains disproportionately low among culturally and linguistically diverse (CALD) women in Australia. Despite national policy efforts, uptake remains low. This study aims to bridge the gap between policy and lived experience by synthesising evidence and capturing perspectives to understand why screening remains inaccessible - and how it can be transformed.

**Methods:** A narrative review of peer-reviewed and grey literature (2015–2024) was conducted to identify structural and cultural barriers affecting cervical screening participation in CALD populations. Key themes were mapped against the socio-ecological model. In parallel, semi-structured interviews were conducted with eight multicultural health workers and cultural advisors across South Australia. Thematic analysis was used to identify recurring patterns, community insights, and practice-level solutions. Ethics approval was granted by the University of South Australia Human Research Ethics Committee.

**Results:** Literature consistently reported language barriers, lack of culturally safe information, and limited awareness of self-collection options. Interviewees deepened this with lived insight, describing mistrust, stigma, and fear, especially among women from refugee and faith-based communities. They proposed actionable strategies: co-designed messaging, partnerships with cultural leaders, and embedding screening education into women’s community spaces. Importantly, the disconnect between national messaging and local realities was seen as a critical barrier to meaningful engagement.

**Conclusions and Significance/Impact:** This study reveals that closing the cervical screening gap for CALD women requires more than information — it demands cultural trust, community leadership, and policy grounded in real-world experience. By combining evidence with voice, it offers a roadmap for developing inclusive, community-led screening programs. The findings advance translational public health by demonstrating how innovation begins with listening — and leads to systems that truly work for all.

## INTRODUCTION

Cervical cancer continues to pose a significant public health challenge in Australia, despite marked improvements in prevention and early detection strategies. The introduction of the Human Papillomavirus (HPV) vaccination program in 2007 and the shift from Pap smears to primary HPV testing in 2017 have significantly reduced cervical cancer incidence and improved the accuracy of early detection (Cancer Australia, 2024). Nevertheless, disparities persist in screening participation and outcomes, particularly among Culturally and Linguistically Diverse (CALD) women, who experience lower screening uptake and higher barriers to timely diagnosis compared with the general population (Australian Institute of Health and Welfare [AIHW], 2024). Understanding and addressing these disparities is essential for achieving equitable health outcomes and aligning with Australia’s broader goal of eliminating cervical cancer as a public health problem by 2035 (Department of Health and Aged Care, 2024).

### Epidemiological Context

The National Cervical Screening Program (NCSP) invites individuals aged 25 to 74 to undergo cervical screening every five years using HPV testing, aiming to detect high-risk HPV infections that may lead to cervical cancer (Department of Health and Aged Care, 2024). Data from the AIHW show that although national screening participation has generally remained above 50% for the target population, CALD women, especially those who are recent migrants or refugees, consistently report lower participation rates, sometimes as low as 30% (AIHW, 2024). This disparity reflects not only gaps in access but also social, cultural, and informational barriers that inhibit engagement with preventive health services.

### Barriers to Screening

A wide range of structural, cultural, and individual-level barriers impede cervical screening among CALD women. Structural barriers include geographic limitations, limited availability of female practitioners, and difficulties navigating healthcare systems (Chandrakumar et al., 2022). Language differences and a lack of culturally appropriate health information further hinder understanding of the purpose and process of cervical screening (Schoenborn et al., 2020). At the social and cultural level, stigma related to sexual health, fear of judgment, and mistrust of healthcare systems have been identified as key barriers, particularly for women from refugee and faith-based communities (Forbes et al., 2019). Such factors are compounded by limited health literacy and varying cultural understandings of preventive health, which may reduce motivation or willingness to participate in screening programs.

### Enablers of Screening

Conversely, research indicates that specific interventions can successfully increase participation among CALD women. These include community-based and culturally tailored approaches, education delivered by trusted community leaders, and the integration of health messaging into familiar community settings (Zhang et al., 2021). Technological and procedural innovations, such as self-collection HPV tests, have also been shown to overcome barriers related to modesty, privacy, and logistical constraints (McBride et al., 2021). Evidence suggests that combining structural supports with culturally sensitive engagement strategies yields the most significant improvements in participation rates, emphasizing the need for multi-level interventions.

### Policy and Programmatic Context

Recognizing these disparities, the Australian government has implemented policy and programmatic initiatives aimed at enhancing cervical screening uptake among under-screened populations. The NCSP now allows self-collection of samples for individuals who are overdue for screening or who prefer this method, providing an alternative to clinician-collected tests and addressing barriers related to modesty or discomfort (Department of Health and Aged Care, 2024). National strategies to eliminate cervical cancer, including the National Cervical Screening Program and HPV vaccination programs, aim to reduce incidence, mortality, and inequities by setting measurable targets for vaccination, screening, and treatment (Department of Health and Aged Care, 2024). However, these programs have not always accounted for the nuanced cultural and social factors influencing CALD women’s engagement, highlighting the need for community-informed approaches.

### Rationale for the Study

Despite policy advancements and the introduction of self-collection options, knowledge gaps remain regarding the lived experiences and perspectives of CALD women, as well as the insights of health professionals supporting them. Many existing studies focus on quantitative measures, such as participation rates or demographic correlations, but fail to explore the underlying social, cultural, and systemic factors that shape behaviour. This study addresses this gap by combining a narrative review of peer-reviewed and grey literature with semi-structured interviews of multicultural health workers and cultural advisors in South Australia. By integrating evidence synthesis with qualitative insights, this study seeks to identify both barriers and enablers to cervical screening and develop actionable strategies to improve access and participation among CALD women. The findings are expected to inform culturally sensitive, evidence-based, and community-led interventions that align national policy objectives with the lived realities of diverse communities.

In summary, understanding the multifaceted barriers to cervical screening among CALD women is crucial for achieving equitable health outcomes and progressing toward the elimination of cervical cancer in Australia. This study aims to bridge the gap between policy and practice by combining literature evidence with the perspectives of those working closely with CALD communities, providing actionable insights for more inclusive and effective public health programs.

## METHODS

### Study Design

This study adopted a **mixed-methods design**, integrating a **narrative literature review** with **qualitative interviews** to explore the barriers and enablers to cervical cancer screening among culturally and linguistically diverse (CALD) women in Australia. A mixed-methods approach was selected to provide both breadth and depth of understanding: the literature review allowed for mapping of structural and systemic issues, while qualitative interviews captured lived experiences and culturally grounded perspectives that may not be represented in published sources. Integration of findings followed a **convergent design**, whereby insights from both components were synthesized and compared during the analysis phase.

### Literature Review

#### Search Strategy

A narrative review of peer-reviewed and grey literature was conducted to identify existing evidence on cervical cancer screening participation among CALD women in Australia. Databases searched included **PubMed, Scopus, Web of Science, and CINAHL**. Grey literature was identified through Google Scholar, government health websites (e.g., Australian Department of Health, Cancer Council Australia), and reports from non-governmental organisations.

Search terms combined keywords and Medical Subject Headings (MeSH), for example:

- *“cervical cancer screening”* OR *“Pap smear”* OR *“HPV self-collection”*
- AND *“culturally and linguistically diverse”* OR *“CALD”* OR *“migrant”* OR *“refugee”*
- AND *“Australia”*

The search was limited to articles published between **January 2015 and December 2024**, reflecting contemporary screening practices since the introduction of HPV vaccination and self-collection pathways.

#### Inclusion and Exclusion Criteria

Studies were included if they:

1. Focused on cervical cancer screening behaviours or participation among CALD women in Australia.
2. Reported on barriers, enablers, or interventions.
3. Were published in English.

Exclusion criteria included studies that:

1. Focused solely on non-Australian populations without relevance to the local context.
2. Were editorials or opinion pieces without empirical data.
3. Addressed broader cancer screening behaviours without cervical-specific findings.

#### Data Extraction and Synthesis

Titles and abstracts were screened by the primary author (DE), followed by full-text review of eligible studies. Key data extracted included: study design, population characteristics, type of barrier/enabler identified, and recommendations. Findings were mapped against the **socio-ecological model** (individual, interpersonal, organisational, community, and policy levels) to enable structured analysis.

### Qualitative Interviews

#### Participants

Semi-structured interviews were conducted with **eight multicultural health workers and cultural advisors** based in South Australia. These participants were recruited using **purposive sampling** to ensure representation from a range of cultural and community backgrounds, including those working with migrant and refugee populations. Participants were chosen for their professional and community-based roles, which provided them with insights into the health behaviours and needs of CALD women.

Eligibility criteria included:

- Aged 18 years or older.
- Currently working or volunteering in roles related to health promotion, community engagement, or service provision with CALD populations.
- Willing to participate in an interview conducted in English. Recruitment

#### Recruitment

occurred through professional networks, community health organisations, and word-of-mouth referrals. Potential participants were approached via email or phone and provided with an **information sheet** outlining the study aims, ethical considerations, and their rights as participants.

#### Data Collection

Interviews were conducted between **March and May 2024**, either in person or via secure online video conferencing platforms (Zoom or Microsoft Teams). Each interview lasted between **45–60 minutes**. A semi-structured interview guide was used, with questions focused on:

- Perceived barriers to cervical cancer screening among CALD women.
- Cultural beliefs and stigma related to women’s health and screening.
- Awareness and acceptance of self-collection options.
- Recommendations for culturally safe interventions and policy improvements.

All interviews were audio-recorded with participant consent and transcribed verbatim. Identifying details were removed during transcription to preserve confidentiality.

### Data Analysis

#### Literature Review

Extracted data were thematically synthesised using the socio-ecological model as an analytical framework. Findings were categorised into levels (e.g., individual: language and health literacy barriers; policy: gaps in national communication strategies).

#### Qualitative Data

Interview transcripts were imported into **NVivo 14** (QSR International) for coding and analysis. A **thematic analysis** approach, guided by Braun and Clarke’s six-phase framework, was employed:

1. Familiarisation with the data.
2. Initial coding of key concepts.
3. Searching for themes across interviews.
4. Reviewing themes in relation to the coded extracts and overall dataset.
5. Defining and naming themes.
6. Producing a narrative synthesis.

Themes emerging from the qualitative data were compared with the literature review findings to identify areas of convergence (where both data sources aligned) and divergence (where interviews revealed new or overlooked insights).

#### Ethical Considerations

Ethical approval for this study was obtained from the **University of South Australia Human Research Ethics Committee**. All participants provided **written informed consent** prior to participation. Participants were assured of their right to withdraw at any time without consequence, and their confidentiality was maintained throughout the study. Data were stored securely on password-protected servers accessible only to the research team.

#### Rigor and Trustworthiness

To enhance rigor, multiple strategies were applied:

- **Triangulation** of data sources (literature + interviews).
- **Peer debriefing** with academic supervisors to refine coding and interpretation.
- **Member checking**, whereby a summary of findings was returned to interviewees for feedback to ensure accuracy.
- **A reflexive journal** was kept by the researcher to document assumptions, decisions, and reflections throughout the study process.

## RESULTS

### Literature Review Findings

A total of **32 sources** (24 peer-reviewed articles and 8 grey literature reports) were included after screening. These sources consistently highlighted **multilevel barriers and enablers** to cervical cancer screening among CALD women in Australia. Findings are summarised below and mapped to the socio-ecological model.

### Individual-Level Barriers

- **Language and health literacy:** Limited English proficiency emerged as a dominant barrier, with many women unable to navigate appointment booking systems, understand medical terminology, or interpret written information (Butler et al., 2020; Cancer Council Victoria, 2022).
- **Knowledge gaps:** Awareness of HPV vaccination and self-collection options was particularly low among older migrant women and those from refugee backgrounds (Sultana et al., 2021).
- **Fear and embarrassment:** Fear of pain, discomfort, or cancer diagnosis, alongside embarrassment in discussing gynaecological health, deterred participation.

### Interpersonal and Community-Level Barriers

- **Cultural norms and stigma:** In some communities, cervical screening was perceived as linked to promiscuity or inappropriate for unmarried women (Musa et al., 2019).
- **Mistrust of the health system:** Negative past experiences or perceived discrimination within healthcare settings reduced willingness to engage.
- **Influence of family and community leaders:** Male partners’ disapproval and limited advocacy from cultural leaders were cited as key deterrents.

### Organisational and Systemic Barriers

- **Inadequate cultural safety:** Studies reported a lack of interpreters, limited training for health professionals in cross-cultural communication, and insufficient use of bilingual health workers (Mishra et al., 2022).
- **Service accessibility:** Barriers included inconvenient clinic hours, transport difficulties, and limited childcare support.

### Policy-Level Barriers

- **Disconnect between policy and practice:** Despite national strategies promoting self-collection, implementation at the community level remained inconsistent (Canfell et al., 2018).
- **Underrepresentation in health promotion:** CALD communities were not adequately included in co-design processes for national awareness campaigns.

### Reported Enablers

Across the literature, several enablers were identified:

- **Availability of female practitioners** improved uptake.
- **Community-based education sessions** delivered in native languages showed positive impacts.
- **HPV self-collection kits** were widely reported as an acceptable alternative, particularly when offered in trusted community settings.

### Qualitative Interview Findings

Interviews with eight multicultural health workers and cultural advisors generated **four overarching themes** with subthemes, reflecting the lived realities of CALD women.

#### Theme 1: Language and Health Literacy as Persistent Barriers

Participants stressed that language barriers extended beyond translation. Many women could not interpret medical jargon or navigate complex referral pathways. As one participant explained:

> *“Even if we translate the leaflet, the way health is explained doesn’t connect with how women in our community understand the body. It’s more than words—it’s worldview.”* (Participant 4)

#### Theme 2: Stigma, Fear, and Cultural Beliefs

Interviewees described strong stigma attached to cervical cancer and its screening. In some communities, screening was associated with promiscuity or moral judgement.

> *“Some women feel that doing this test is admitting to being sexually active, and in their culture, that is judged very harshly.”* (Participant 7)

Fear of results, fatalistic views of cancer, and cultural taboos around discussing reproductive health reinforced avoidance.

#### Theme 3: Trust and Relationships Matter

Participants highlighted mistrust of mainstream health services, particularly among refugee-background women who had experienced systemic discrimination. Building trust through community-based initiatives and bilingual health workers was considered critical.

> *“Women don’t go because they don’t trust the system will treat them with dignity. But if it’s the nurse from their own community, they’ll listen.”* (Participant 3)

#### Theme 4: Pathways to Culturally Safe Screening

Several enablers were identified by participants:

- **Co-designed messaging:** Campaigns developed with cultural leaders, faith-based organisations, and women’s groups.
- **Use of trusted spaces:** Embedding self-collection options in community centres, libraries, and women’s groups rather than clinics.
- **Female practitioners and interpreters:** Ensuring women felt comfortable during screening.
- **Education through community leaders:** Leveraging cultural and religious leaders to endorse participation.

### Integration of Findings

When combined, the literature and interviews revealed both **overlap** and **new insights**.

- Overlap: Both sources identified language barriers, stigma, and the importance of female practitioners.
- New insights: Interviews highlighted **deep mistrust**, the role of **fatalistic beliefs**, and the importance of **community trust**—themes less visible in published literature.

The integration suggests that while policy has advanced (e.g., through HPV self-collection), **structural and cultural barriers remain deeply entrenched**, requiring community-led solutions to bridge the gap between national strategy and local reality.

## DISCUSSION

This mixed methods study examined the barriers and enablers to cervical cancer screening among culturally and linguistically diverse (CALD) women in Australia by integrating evidence from a narrative review with perspectives from multicultural health workers and cultural advisors. Findings revealed a complex interplay of structural, cultural, and interpersonal factors that continue to limit screening participation, despite advances in prevention and policy innovation. Importantly, the study highlights the gap between national strategies and the lived realities of CALD communities, underscoring the need for culturally responsive, community-driven approaches.

### Interpretation of Key Findings

#### Barriers across multiple levels

Consistent with previous studies (Butler et al., 2020; Sultana et al., 2021), our findings confirmed that language barriers, limited awareness of self-collection options, and fear or stigma surrounding cervical cancer remain significant deterrents. Health literacy challenges were not only about translating materials but about ensuring that biomedical concepts are explained in ways that align with cultural worldviews. This aligns with international evidence showing that cultural explanatory models of disease often differ from biomedical explanations, and mismatches can lead to disengagement with screening (Musa et al., 2019).

At the interpersonal level, stigma surrounding sexual health, particularly for unmarried or younger women, emerged strongly in both the literature and interviews. The association between screening and promiscuity has been reported in migrant and refugee communities across Europe and North America, suggesting a global challenge (Fang et al., 2018). Our findings confirm that these perceptions remain salient in Australia, reinforcing the importance of culturally sensitive education campaigns.

At the systemic and policy level, participants described a disconnect between national screening strategies and community-level implementation. While self-collection has been promoted as a game-changer (Canfell et al., 2018), awareness and accessibility remain low in CALD communities. This highlights a broader issue in public health: policies that assume equitable uptake often fail without targeted outreach to marginalised groups.

#### The role of trust and community leadership

A distinctive contribution of this study lies in the identification of trust as a central enabler. While the literature points to mistrust as a barrier, interviews illustrated how trust can be built through community leadership, cultural advisors, and bilingual health workers. Participants consistently noted that women are more willing to engage when screening is endorsed by respected figures in their cultural or religious communities. This finding resonates with community-based participatory research (CBPR) approaches, which have demonstrated effectiveness in addressing health inequities by fostering trust and co-designing interventions with communities (Israel et al., 2017).

#### HPV self-collection as an opportunity

Both the literature and interviews underscored the potential of HPV self-collection kits to overcome barriers of embarrassment, modesty, and time constraints. However, uptake remains uneven because distribution strategies are not fully tailored to CALD women. Embedding self-collection into trusted community settings—such as cultural centres, libraries, or women’s groups—emerged as a practical pathway. This aligns with findings from the Compass trial and other implementation studies that highlight the importance of delivery settings in determining uptake (Sultana et al., 2021).

## IMPLICATIONS FOR POLICY AND PRACTICE

### Moving from policy rhetoric to local realities

Despite national policy efforts to increase participation, CALD women remain underrepresented in screening programs. This study suggests that strategies cannot rely solely on universal campaigns; rather, they must address the socio-cultural and structural barriers specific to CALD populations. Co-designed health promotion initiatives, delivered in collaboration with community leaders, could increase cultural resonance and trust.

### Prioritising cultural safety in health services

The findings reinforce the need for cultural safety within clinical practice. This goes beyond providing interpreters to include training health professionals in cross-cultural communication, actively recruiting bilingual practitioners, and addressing structural discrimination within health systems. The principle of cultural safety, first developed in Indigenous health contexts, is increasingly recognised as central to equitable health care delivery for CALD populations (Curtis et al., 2019).

### Harnessing community networks

Community organisations, religious institutions, and multicultural health services represent underutilised assets in cervical cancer prevention. Investing in partnerships with these networks could significantly improve outreach. Programs that integrate screening education into existing community events—such as women’s groups, cultural festivals, or religious gatherings—may normalise participation and reduce stigma.

### Enhancing the roll-out of self-collection

Self-collection holds promise for CALD women, but equitable access requires targeted distribution strategies. For example, mailing kits may not be effective for women with unstable housing or limited literacy. Offering kits through community health workers, at trusted community sites, or during home visits could help ensure that innovations reach the populations most in need.

### Comparison with International Literature

The barriers identified in this study mirror those reported internationally. Studies in the United States, United Kingdom, and Canada consistently describe language barriers, stigma, mistrust, and lack of culturally appropriate care as obstacles to screening among immigrant and refugee women (Fang et al., 2018; Marlow et al., 2019). The emphasis on self-collection as an enabler also reflects global findings. However, our study adds nuance by highlighting how CALD women in Australia perceive the disconnect between national messaging and their lived experiences.

### Strengths and Limitations

A key strength of this study is its mixed-methods design, combining a narrative review with qualitative interviews to capture both the evidence base and lived perspectives. This integration strengthens the validity of findings and ensures that recommendations are grounded in both data and community voice.

However, several limitations should be noted. First, the number of interviews was modest (n=8), which may limit generalisability. Participants were multicultural health workers and advisors rather than CALD women themselves, meaning insights were indirect. Second, while the narrative review included both peer-reviewed and grey literature, it was not a systematic review and may not have captured all relevant studies. Finally, findings are situated in the Australian context, though they may hold relevance for other multicultural settings.

### Future Directions

Future research should directly engage CALD women across different cultural groups to capture their perspectives firsthand. Co-designed intervention studies, particularly those testing community-based models of self-collection, are urgently needed. In addition, longitudinal research could explore how trust-building initiatives influence screening uptake over time.

## CONCLUSION

This study highlights that improving cervical cancer screening among CALD women in Australia requires more than policy innovation; it requires **trust, cultural safety, and community leadership**. While barriers such as language, stigma, and systemic exclusion remain entrenched, enablers including self-collection, female practitioners, and co-designed education offer pathways forward. By bridging the gap between national strategies and community realities, policymakers and practitioners can move toward a more inclusive screening program that truly serves all women.

## Data Availability

All data produced in the present study are available upon reasonable request to the authors

